# Genetic admixture predictors of fetal alcohol spectrum disorders (FASD) in the South African Cape Coloured population

**DOI:** 10.1101/2024.03.31.24305130

**Authors:** R. Colin Carter, Zikun Yang, Tugba Akkaya-Hocagil, Sandra W. Jacobson, Joseph L. Jacobson, Neil C. Dodge, H. Eugene Hoyme, Steven H. Zeisel, Ernesta M. Meintjes, Caghan Kizil, Giuseppe Tosto

## Abstract

Ancestrally admixed populations are underrepresented in genetic studies of complex diseases, which are still dominated by European-descent populations. This is relevant not only from a representation standpoint but also because of admixed populations’ unique features, including being enriched for rare variants, for which effect sizes are disproportionately larger than common polymorphisms. Furthermore, results from these populations may be generalizable to other populations. The South African Cape Coloured (SACC) population is genetically admixed, with one of the highest prevalences of fetal alcohol spectrum disorders (FASD) worldwide. We profiled its admixture and examined associations between ancestry profiles and FASD outcomes using two longitudinal birth cohorts (*N*=308 mothers, 280 children) designed to examine effects of prenatal alcohol exposure on development. Participants were genotyped via MEGA-ex array to capture common and rare variants. Rare variants were overrepresented in our SACC cohorts, with numerous polymorphisms being monomorphic in other reference populations (e.g., ∼30,000 and ∼221,000 variants in gnomAD European and Asian populations, respectively). The cohorts showed global African (51%; Bantu and San); European (26%; Northern/Western); South Asian (18%); and East Asian (5%; largely Southern regions) ancestries. The cohorts exhibited high rates of homozygosity (6%), with regions of homozygosity harboring more deleterious variants when lying within African local-ancestry genomic segments. Both maternal and child ancestry profiles were associated with FASD risk and altered severity of prenatal alcohol exposure-related cognitive deficits in the child. Our findings indicate that the SACC population may be a valuable asset to identify novel disease-associated genetic loci for FASD and other diseases.

## INTRODUCTION

Fetal alcohol spectrum disorders (FASD) are the most common preventable cause of developmental disabilities worldwide, with prevalence estimates of 2-5% in school-aged children in the US and even higher prevalence in endemic communities (1–3). Fetal alcohol syndrome (FAS), the most severe form of FASD, is characterized by neurodevelopmental deficits and specific facial dysmorphology, small head circumference, and growth restriction (4–6). Teratogenic effects of alcohol have been reported in every organ system (5–9). The intricate etiology of FASD underscores a compelling need to explore the genetic landscape contributing to its development. Children with prenatal alcohol exposure (PAE) display remarkable variability in the severity of neurocognitive deficits not explained by dose and timing of exposure alone. However, few genetic determinants of FASD risk have been identified, possibly due to prior studies being limited to candidate genes (mainly alcohol metabolism genes (10)), employing small sample sizes, and examination of non-admixed populations (11). The roles of gene-environment interactions alongside ancestry-related genetic regulatory mechanisms may further complicate the genetic architecture of FASD and susceptibility. The South African Cape Coloured (SACC) community, renowned for its rich tapestry of African, European, South Asian, and East Asian ancestries, presents a unique opportunity to investigate the genetic susceptibility factors of FASD. There is growing interest in investigating admixed populations (i.e., those comprising individuals whose genome is the result of interbreeding between distinct ancestral populations) and their phenotypic presentations. Previous studies have led to key insights into the genetic architecture of rare and common diseases by leveraging ancestry-specific disease-associated variants/genes/pathways or methods, i.e., admixture mapping (12). Admixed populations are known to be enriched for rare variants, leading to increased discovery power. Compared with common variants, rare variants are more likely to have higher risk- and protection-associations in complex diseases. Identification of the *APOE3* Christchurch mutation in Alzheimer’s disease, which was found in a Colombian admixed population, typifies the recent discovery of novel rare variants in admixed populations (13). Disease-associated alleles identified in one population, even if absent in others, may still be used to develop therapeutics that work in all people. For example, alleles in the *PCSK9* gene are associated with a 40% reduction in low density lipoprotein in an African American cohort (14). Although these variants are extremely rare in Europeans, drugs based on this discovery are effective in most individuals.

The South African Cape Coloured (SACC) population has extensive genetic admixture that reflects a complex history of colonialism, apartheid-era enforced segregation, and three decades of democracy. Although race is a social construct, geopolitical and institutional practices and policies over the last three centuries have heavily influenced the genetic admixture of the SACC population. Previous studies have demonstrated genetic ancestry from the African Khoe-San, African Bantu-speaking, European, South Asian, and East Asian populations (15–19). The SACC population may be particularly well-suited to genetic association studies for complex diseases, such FASD. Notably, the prevalence of FASD in the SACC population is among the highest in the world, affecting 13.6-30.6% of school-aged children (20). To our knowledge, the only study that has examined genetic risk factors for FASD in the SACC population was a case control study focusing on the mothers’ alcohol dehydrogenase-1B, *ADH1B,* genotype, which has been associated with altered fetal vulnerability in other populations (10, 21); this SACC cohort study cohort showed a higher frequency of the *rs1229984* allele, which is associated with more efficient alcohol metabolism, among mothers of unexposed control children vs. mothers of children with FASD (22).

Here, we employed genome-wide genotyping data from two SACC prospective longitudinal birth cohorts in Cape Town, South Africa, designed to examine the developmental effects of PAE to (a) describe the genetic admixture of SACC population and (b) examine whether ancestry is associated with FASD risk.

## METHODS

### Cohorts’ description

Data are presented from two prospective longitudinal cohorts designed to examine the developmental effects of PAE: one recruited from 1999-2002 (“Cohort 1”) (9, 23); the other from 2001- 2015 (“Cohort 2”)(24, 25). Alcohol consumption at recruitment was ascertained in timeline follow-back interviews (26, 27). Women were invited to participate if they averaged at least 1.0 oz (30 mL) absolute alcohol (AA)/day (≈1.67 standard drinks) or reported binge drinking (based on National Institute on Alcohol Abuse and Alcoholism (NIAAA) binge definition at the time of recruitment: ≥2.5 oz (75 mL) AA/drinking occasion for Cohort 1; (≥2.0 oz (60 mL) AA/ occasion for Cohort 2). Women who abstained or drank only minimally (with no binge episodes) were invited to participate as controls.

Exclusion criteria at recruitment included maternal age <18 years, HIV infection, multiple gestation pregnancy, and pharmacologic treatment for medical conditions. Infant exclusion criteria included major chromosome anomalies, recognizable malformation syndromes (other than FASD), neural tube defects, very low birthweight (<1500 g), and extreme prematurity (gestation <32 weeks). DNA samples were unavailable for both mother and child for 18 mother-child dyads with heavy exposure and 13 controls in Cohort 1; 7 mother-child dyads with heavy exposure and 7 controls in Cohort 2. Nine participants failed initial quality control procedures). The final cohorts for the analyses presented here included 128 mother-infant dyads (76 with heavy PAE, 52 controls) for Cohort 1; 200 mother-infant dyads (120 with heavy PAE, 80 controls) for Cohort 2. 32 heavy drinking mothers in Cohort 2 received dietary choline supplementation as part of a pilot study we were conducting (28); their children were not genotyped and were not included in analyses for which child phenotype was the outcome.

Each woman provided informed consent, and interviews were conducted in the mother’s preferred language (Afrikaans or English). Women reporting alcohol or drug use were counseled by our staff about the risks of prenatal alcohol and drug use, were encouraged to stop or reduce their alcohol and/or drug use and were offered referrals to substance use treatment programs.

### Prenatal alcohol exposure and phenotypic characterizations of mothers and children

In timeline follow-back interviews (26) administered at recruitment and mid-pregnancy (Cohort 1) or 4 and 12 weeks after recruitment (Cohort 2), each mother was asked about her alcohol, smoking, and drug use on a day-by-day basis during the past 2 weeks, with recall linked to specific daily activities. Volume was recorded for each type of alcohol beverage consumed and converted to oz AA using weights proposed by Bowman et al. (29). Maternal height was obtained in postnatal study visits for Cohort 1 and prenatal study visits for Cohort 2 by research staff trained by RCC (25). Recognition memory assessments were performed in both cohorts: the California Verbal Learning Test-Children’s version (Pearson Clinical Assessments, USA) was administered at age 10 years in Cohort 1; the Wide Range Assessment of Memory and Learning, 2^nd^ Edition (Pearson Clinical Assessments, USA) was administered at age 7 years for Cohort 2. In 2005, 2009, 2013, and 2016, we held FASD diagnostic clinics, in which all enrolled children were independently examined by expert FASD dysmorphologist HE Hoyme MD and other trained FASD dysmorphologists for growth and FAS-related anomalies. All examiners were blind regarding PAE and prior diagnoses. There was substantial agreement among dysmorphologists on assessments of all dysmorphic features, including philtrum and vermilion ratings based on the lip/philtrum guide (30) and palpebral fissure length. Each case was reviewed in case conferences by HEH, RCC, SWJ, CDM, and JLJ, who jointly determined FASD diagnosis using the Hoyme criteria (5).

### Blood sample collection, DNA extraction, and genotyping

A 10 ml venous EDTA whole blood sample was obtained from each mother and child in Cohort 1 in 2009 at the local diagnostic clinic. For Cohort 2, a 10 ml venous EDTA whole blood sample was obtained from mothers at a prenatal study visit (2011–2015); saliva samples were obtained with oragene- 575 kits (DNAgenotek, Ottawa, Canada) from infants in the 1^st^ year of life. For mothers or children from both clinics missing a venous blood sample (e.g., due to missed visits or failed blood draws), saliva samples were obtained using oragene-500 kits (DNAgenotek, Ottawa, Canada). All samples were stored at -80 degrees C (within 24 hours for blood and within 1 year for saliva (per manufacturer instructions)) and shipped with continuous cold chain until processing for DNA. DNA was isolated using the Maxwell® 16 LEV Blood DNA Kit (Promega, #AS1290, Madison, WI) and stored at - 20°C until genotyping assays were performed. All samples were assayed using the Illumina (San Diego, USA) Infinium Expanded Multi-Ethnic Genotyping Array (MEGA^EX^), which contains >2 million SNPs and covers 65.7% of GWAS catalog SNPs. Genotyped data were cleaned with standard QC measures using PLINK (v1.9). In brief, SNPs were excluded if Hardy-Weinberg equilibrium *p*-value <1-E06 and missing call rate <95%; individuals were removed if genotype missingness ≥2%. Principal component analysis was conducted using *king* software (31) to account for variance in population stratification and to identify unrelated set of samples (defined as kinship <0.09).

### Ancestry inference

We used a local ancestry inference method, ELAI (32), to conduct the chromosome-wise ancestry inference and then computed the genome-wide ancestry dosage for each individual in our cohorts. We used a newly-developed reference database (33), which harmonized two public databases: 1000 Genome (1000G) (34) and the Human Genome Diversity Project (HGDP) (35). Consistent with prior studies in this population (15–19), we conducted supervised admixture analyses utilizing African, European, South Asian, and East Asian as reference population data [African: HGDP Yoruba, Bantu Kena, Mandenka and 1000G Yoruba; European: HGDP French, Tuscan, Bergamo Italian and 1000G Great Britain, CEU (Utah, USA residents with Northern and Western European), Toscani Italian; South Asian HGDP Cambodian, Lahu (ethnic group in Myanmar, Laos, Thailand) and 1000G Bengali (in Bangladesh), Kinh (Vietnamese); East Asian (HGDP Han-Chinese, Northern Han-Chinese and 1000G Southern Han Chinese, Beijing Han Chinese]. PCA analyis of uncorrelated SNPs was then conducted on a combined dataset of 352 unrelated participants in our cohorts and the reference populations listed above to examine similarities between our SACC cohorts participants and the reference populations.

Toward this aim, the software ADMIXTURE was also employed, varying *k* from 4-6, based on prior studies and the ELAI results, using uncorrelated genetic markers in 352 unrelated individuals from our SACC cohorts participants and HGDP and 1000G African, European, South Asian, and East Asian reference populations.

### Runs of Homozygosity (ROH)

ROHs are continuous homozygous stretches of the genome which indicate ancestral relatedness and, especially when long, evidence of recent inbreeding. We estimated ROH employing 296 unrelated mothers from our SACC cohorts using the Plink software (36) and computed the correlation between overall ROH and global ancestry.

We then selected the top 10 ROH regions most frequently mapped across the 296 mothers. We further utilized the results of ELAI (32) and ANNOVAR (37) to determine the number of deleterious variants within a certain local ancestry background (**Supplemental Figure S1**). For each individual, we estimated the local ancestral background within the prioritized ROHs. We then excluded individuals with ancestral switch within those regions (e.g. AFR -> EUR) and classified samples in those carrying a ROH traced back to African ancestry, vs those with ROH lying in non-African genomic segments. A variant was defined as deleterious if annotations from ANNOVAR classified the variants as probably damaging or deleterious (i.e., SIFT, PolyPhen-2 HDIV, PolyPhen-2 HVAR, PROVEN, FATHMM-MKL), indicating that the variant is expected to affect protein function and likely to be deleterious. We tallied the ROH ancestral background and the respective counts of deleterious variants and computed a poisson test to test whether African ROH vs. non-African ROH were enriched for deleterious variants.

### Statistical Analysis

All statistical analyses were conducted using SPSS (v.24; IBM, Armonk, NY, USA) or R (R Foundation, Vienna, Austria). All variables were examined for normality of distribution; oz AA/day were log-transformed due to skewness (>3.0). Independent samples *t*-tests were used to compare continuous outcomes between groups (e.g., heavy drinking vs. control women); chi-square/Fisher’s exact test for categorical outcomes. Given the widely held assumption that Cape Malay (Muslim- identified SACC) individuals are genetically distinct from other SACC individuals, we compared ELAI- estimated ancestry proportions between these groups using independent samples *t*-tests. To examine potential associations between ancestry and maternal outcomes, linear regression models were constructed regressing the given outcome on each ancestry proportion and potential confounders (maternal age, socioeconomic status, and cohort). To examine potential associations between ancestry and child FASD diagnosis, logistic regression models were constructed regressing FAS/PFAS diagnosis vs. non-syndromal on each ancestry proportion, and potential confounders (maternal age, socioeconomic status, cohort, and PAE). To examine ancestry-by-PAE interaction effects, linear regression models were constructed regressing the recognition memory Z-scores on each ancestry proportion, PAE (*ln*(average oz absolute alcohol/day across pregnancy)), a PAE-by-ancestry proportion interaction term, and maternal age and socioeconomic status.

## RESULTS

### Sample characteristics

Women in both cohorts were in their 3^rd^ decade on average, and heavy drinking mothers were older than controls (**Table 1**). Heavy drinking and control mothers completed 8-10 years of school, on average, with heavy drinking women averaging fewer years of school than controls. Heavy drinking mothers in both cohorts exhibited a weekend-binge pattern of drinking, averaging 7.0 standard drinks/occasion on 1.4 days per week in Cohort 1 and 6.7 standard drinks/occasion on 1.4 days per week in Cohort 2. Most control women abstained from drinking during pregnancy, with 12 women reporting infrequent, light consumption (no binges (i.e, <4 standard drinks/occasion), with drinking occasions <2/month for 10, <1/week for 2). Cigarette smoking was more common among heavy drinking women than controls in Cohort 1, but the number of cigarettes smoked per day was similarly low in both groups. 14.5% of heavy drinking women vs. 3.8 % of controls in Cohort 1 and 24.2% of heavy drinking women vs. 10.0 % of controls in Cohort 2 reported prenatal marijuana use. 11.7% vs. 15.0% of control women in Cohort 2 reported prenatal methamphetamine use; no women in Cohort 1 reported methamphetamine use, as this was uncommon in the area in the early 2000’s. Methaqualone, cocaine, and opiate use were not reported by any participant. Most women (82.0% in Cohort 1; 86.5% in Cohort 2) delivered at term. 47.4% of exposed children in Cohort 1 and 30.8% of children in Cohort 2 met diagnostic criteria for FAS or partial FAS (PFAS).

**Table 1.**
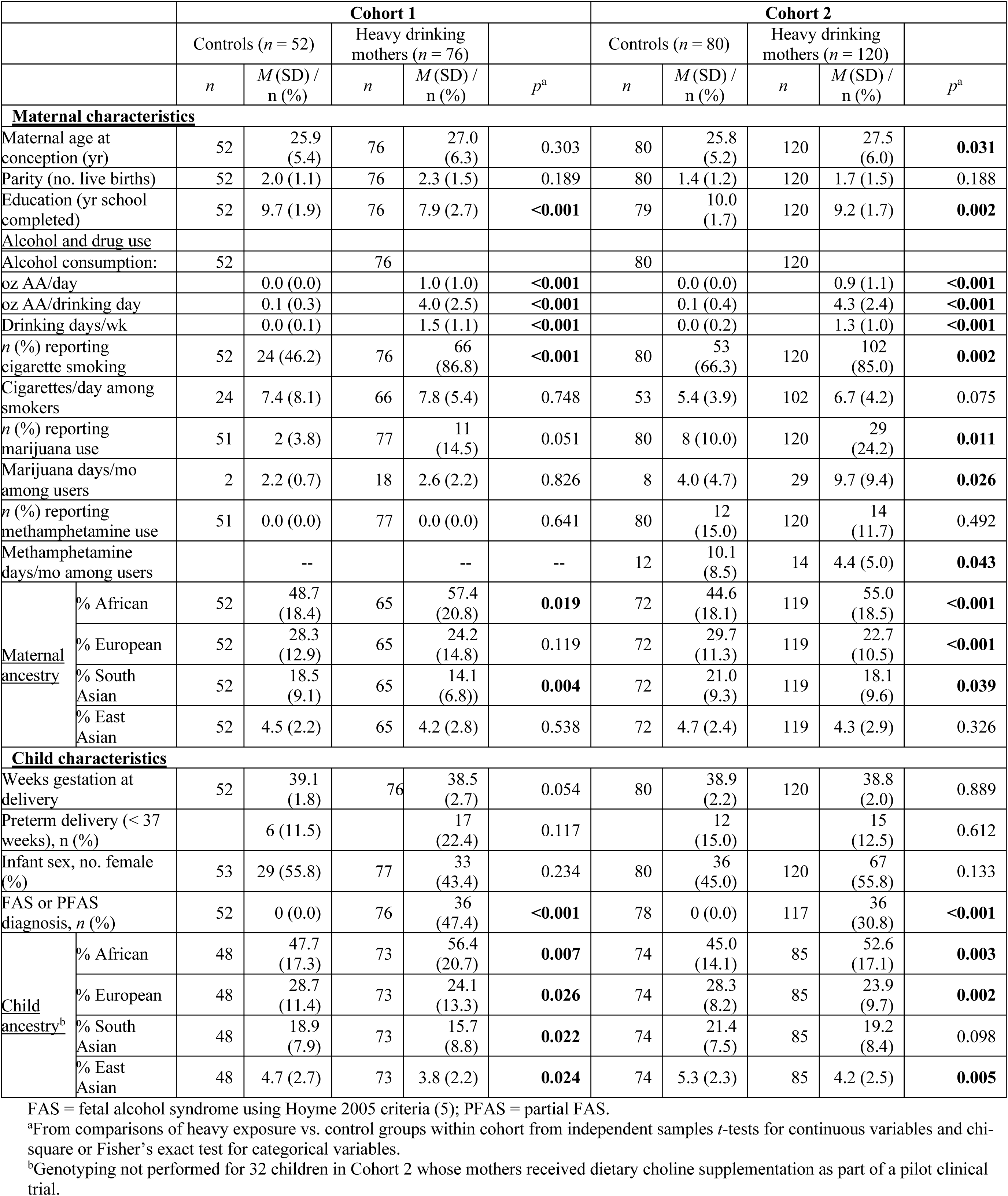
Sample characteristics.

**Table 2.**
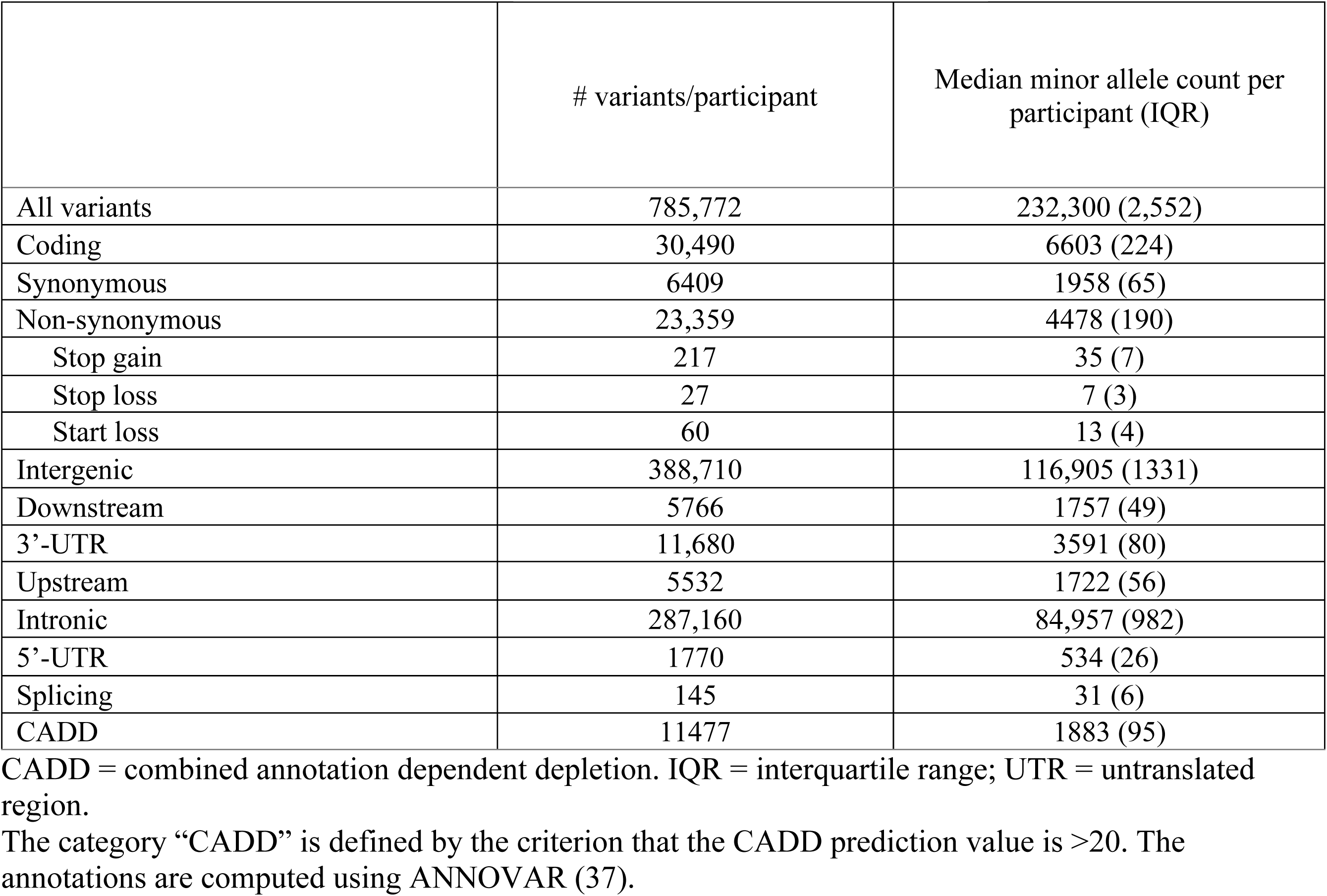
Number of variants discovered in genotyping 588 SACC participants.

### Population structure

After quality control, 785,772 variants remained. Among those, we annotated 6,409 synonymous coding variants, and 23,359 non-synonymous variants, with 304 variants being stop-gain, stop-loss, or start-loss. 2.11% were rare variants (minor allele frequency (MAF) < 1%). **Table 3** shows that variants found to be rare in the gnomAD reference superpopulations (38) are common in SACC. Specifically, 29,847 variants that were monomorphic in gnomAD Europeans, were non-monomorphic in SACC, and 26,853 (90%) of these were common in our cohorts. Relatedness was moderate, which was expected given that individuals were recruited from the same geographic area; among mothers, 42 (13.29%) were first-degree relatives, 10 (3.16%) second-degree, and 22 (7.00%) third-degrees.

**Table 3.** Prevalence of rare and common variants in the SACC cohorts vs. reference populations (*N* = 296 unrelated mothers).

|  | Monomorphic | Ultra Rare<br>(MAF<0.001) | Rare<br>(0.001<MAF<0.01) | Common<br>(MAF>0.01) |
| --- | --- | --- | --- | --- |
| SACC cohorts | 2 | 0 | 16,565 | 769,479 |
| European <sup>a</sup> | 29,847 | 105,395 | 46,934 | 603,750 |
| African <sup>a</sup> | 2,455 | 4,399 | 38,098 | 741,091 |
| East Asian <sup>a</sup> | 221,088 | 33,870 | 473,41 | 483,738 |
| SACC variants that are monomorphic in Europeans |  |  |  |  |
| SACC cohorts | 0 | 0 | 2,994 | 26,853 |

SACC = South African Cape Coloured
<sup>a</sup>gnomAD (38).

### Admixture

ELAI-based (32) ancestry estimations demonstrated predominantly African ancestry (51%), followed by European (26%), South Asian (18%), and a smaller proportion of East Asian (5%) ancestries (**Table 1**, **Figure 1A**). Using Admixture (39), we compared uncorrelated genetic markers in 352 unrelated individuals from our SACC cohorts participants to HGDP and 1000G African, European, South Asian, and East Asian reference populations allowing for 4, 5, and 6 groups (**Figure 1B, Supplemental Table S1**). The four-group analysis was consistent with the ELAI estimations. In the five-group analysis, our SACC cohorts participants’ African ancestry clustered mostly with two individuals of San origin in the African HGDP reference population. In the six-group analysis, our

**Figure 1.**
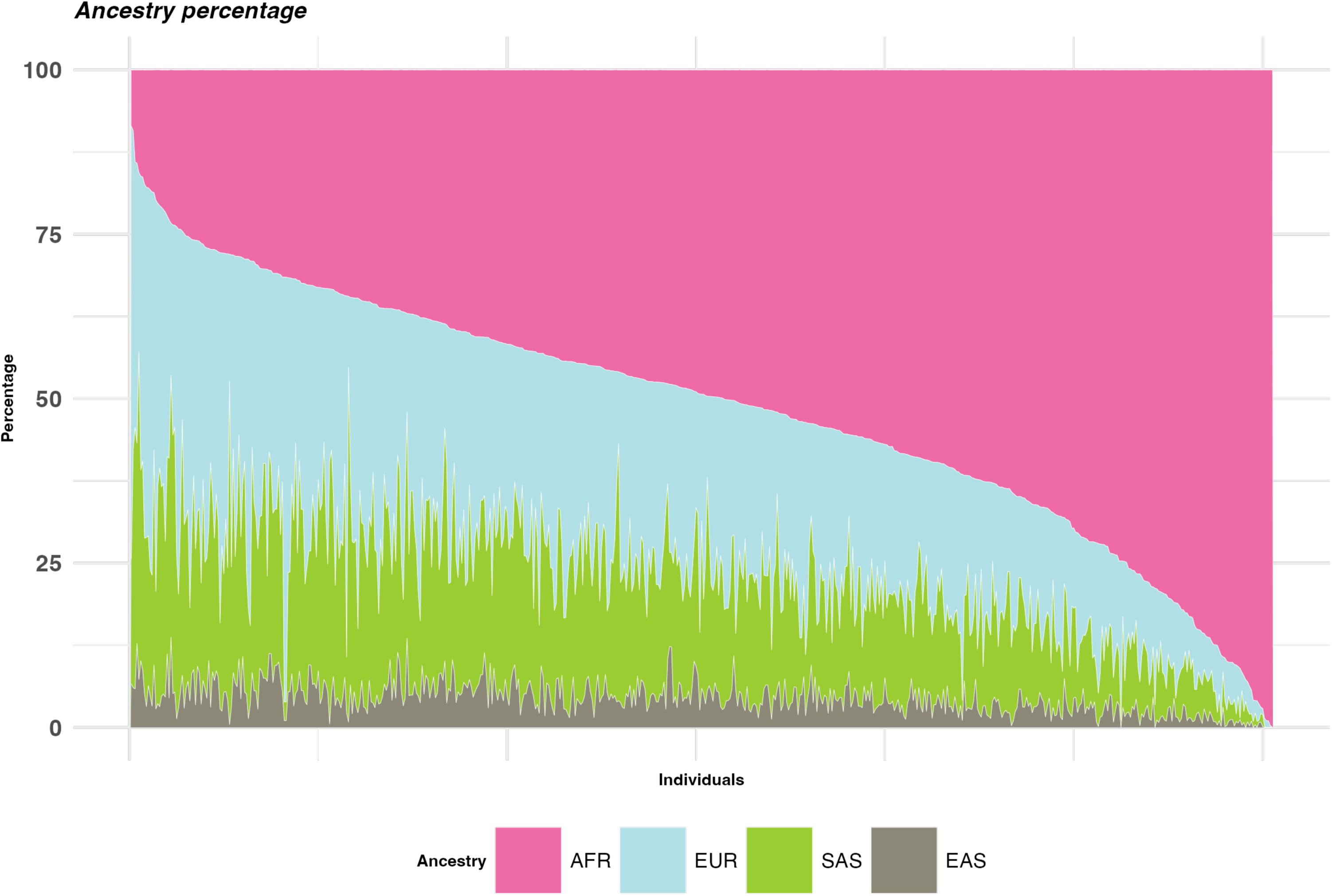
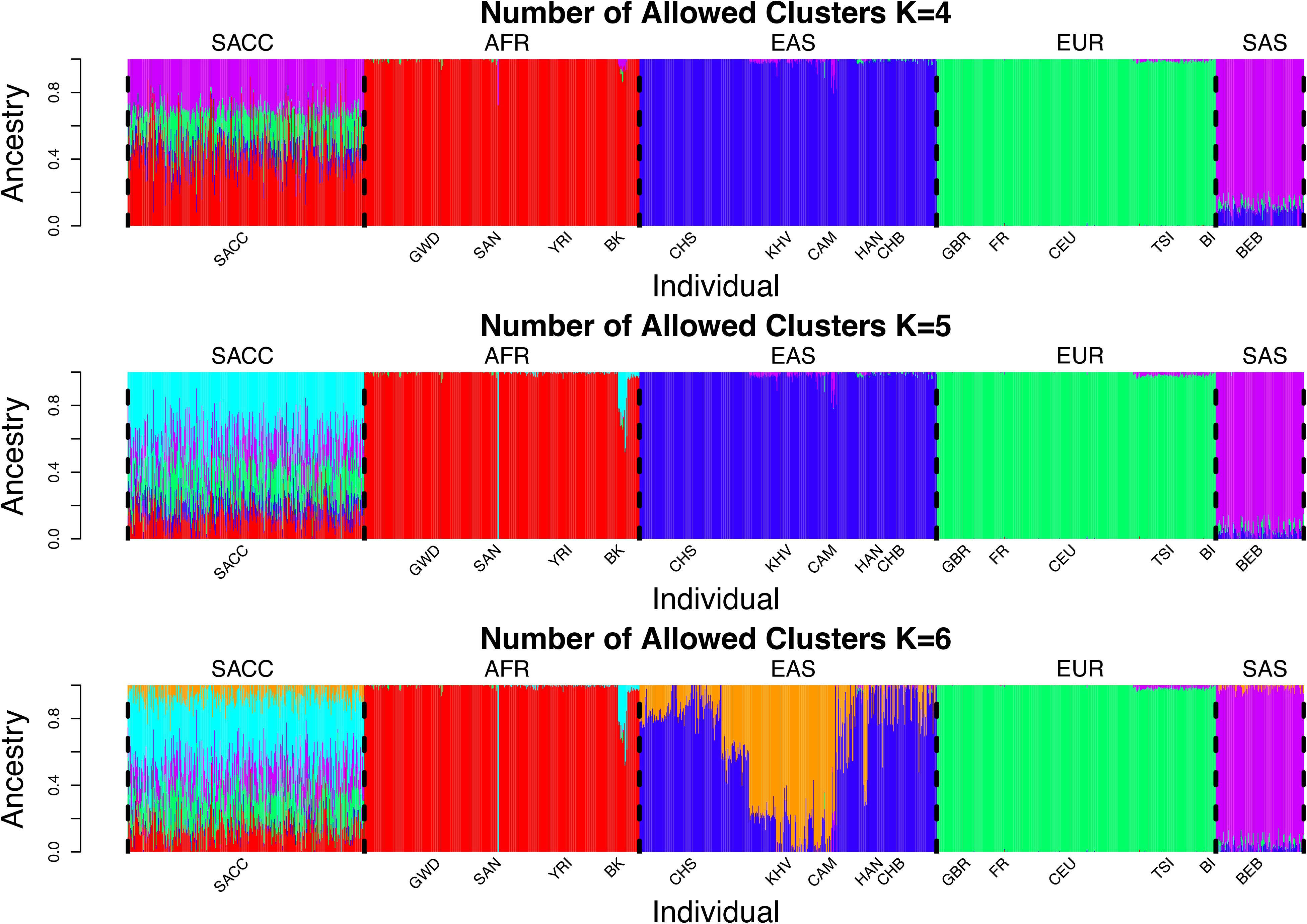
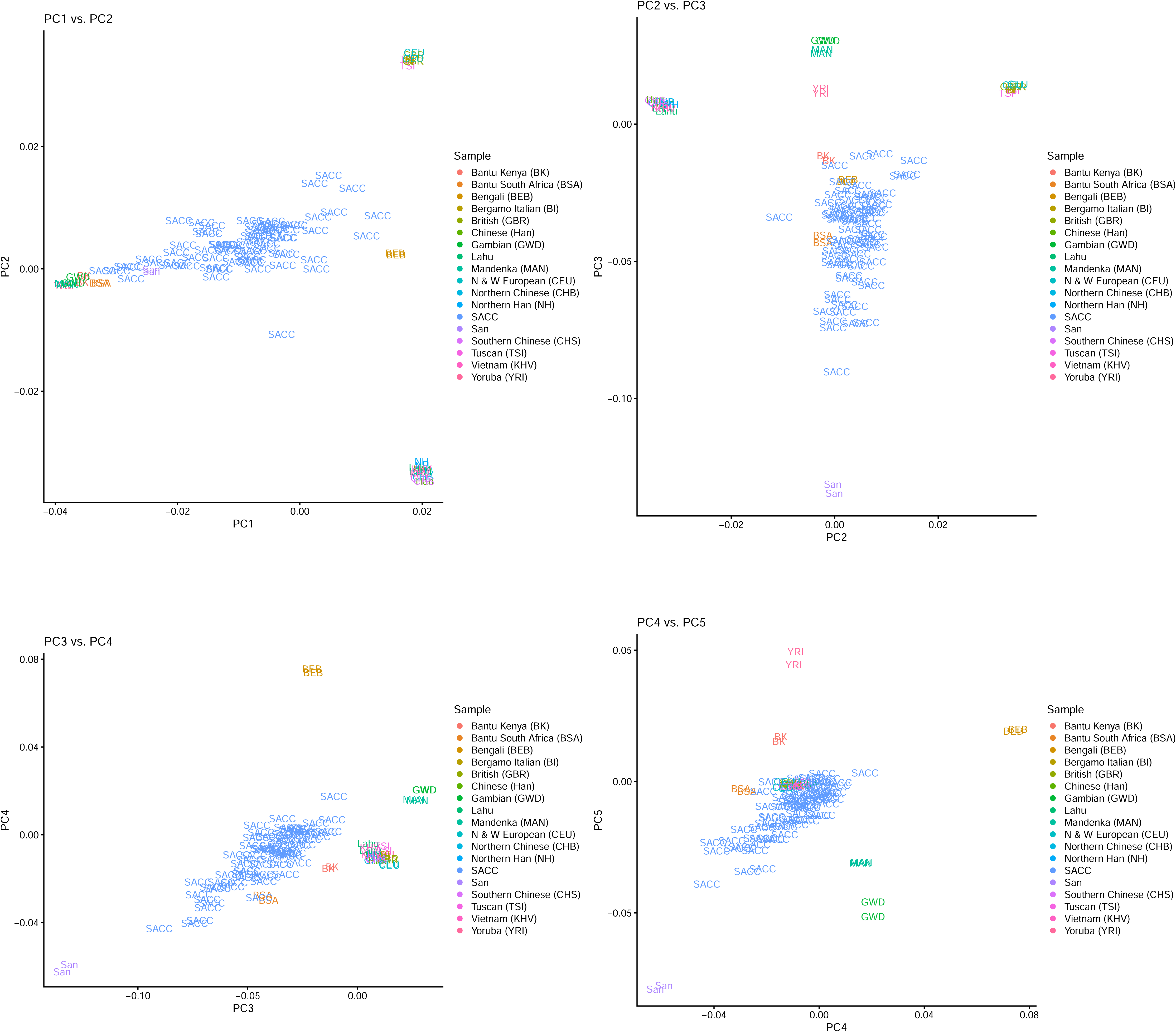
(A) Ancestry proportions (estimated with ELAI (32) method) for all SACC cohort participants and (B) admixture analysis (39) of uncorrelated SNPs in 352 unrelated participants combined with selected African, European, South Asian, and East Asian reference populations allowing for 4, 5, and 6 clusters. (C) PCA analyis of uncorrelated SNPs in a combined dataset of 352 unrelated cohort participants (SACC) and selected reference populations BK = Bantu Kenya; BSA = Bantu South Africa; BEB = Bengali; BI = Bergamo Italian; CAM = Cambodian; CEU = Northern and Western European (from Utah, USA); CHB = Northern Chinese; CHS = Southern Chinese; GBR = British; GWD = Gambian; Han = Chinese; KHV = Vietnamese (Kinh); MAN = Mandenka; NH = Northern Han; SACC = South African Cape Coloured; TSI = Tuscan (Italy); YRI = Yoruba.

SACC cohorts participants’ East Asian ancestry clustered most with the Vietnamese (Kinh; KHV), Lahu (Myanmar, Laos, Thailand), and Cambodian (CAM) reference populations, which are geographically closer to Malaysia and Indonesia, where SACC ancestors are thought to have lived prior to being brought to South Africa. Similarly, in the PCA analyis of uncorrelated SNPs in a combined dataset of 352 unrelated SACC cohorts participants and selected reference populations **(Figure 1C**), SACC participants clustered with San, Bantu Kenyan, Bantu South African, Northern and Western European, Bengali, Kinh (Vietnam), Lahu (Myanmar, Laos, Thailand) and Northern Han Chinese populations.

Cape Malay individuals had lower ELAI-estimated (32) proportions of African ancestry and higher proportions of European, South Asian, and East Asian ancestry proportions (**Supplemental Table S2**.

### Runs of Homozygosity

The homozygosity rate, estimated by the overall run of homozygosity (ROH) from the genotypes of 296 unrelated mothers, was 5.84%. African ancestry proportion was negatively associated with % homozygosity, whereas European, South Asian, and East Asian ancestry proportions were positively associated with % homozygosity (**Supplemental Table S3**). For the ten top ROHs, (**Supplemental Table S4**) the poisson test comparing deleterious variants between African vs non-African ROH segment revealed that African ROH segments harbor a higher number of deleterious variants Ppoisson=0.023).

### Associations between global ancestry and phenotypic outcomes

Linear regression models examining associations ancestry percentages and demographic and phenotypic outcomes are presented in **Table 4**. Relative to the other ancestries, higher maternal African ancestry was associated with lower socioeconomic status (opposite for European ancestry) after control for cohort. Maternal African ancestry was negatively associated with maternal height after control for socioeconomic status and cohort. By contrast, European and South Asian ancestry were positively associated with maternal height. After control for maternal age, socioeconomic status, and cohort, African ancestry percentage was associated with higher levels of alcohol consumption across pregnancy, while European and South Asian ancestry percentages were associated with lesser alcohol consumption.

**Table 4.** Associations between ancestry percentages and demographic and phenotypic outcomes.

|  | AFR | EUR | SAS | EAS |
| --- | --- | --- | --- | --- |
| **<u>Maternal Ancestry (n = 308)</u>** |  |  |  |  |
| Socioeconomic status<sup>a,b</sup> | -0.06*<br>(-0.10, -0.01) | 0.09*<br>(0.02, 0.16) | 0.09<br>(-0.01, 0.19) | 0.04<br>(-0.29, 0.37) |
| Maternal height (cm) <sup>a,c</sup> | -0.07***<br>(-0.11, -0.04) | 0.13***<br>(0.07, 0.18) | 0.08*<br>(-0.00, 0.15) | 0.22†<br>(-0.04, 0.47) |
| Average oz absolute alcohol/day across pregnancy<sup>a,d</sup> | 0.01***<br>(0.00, 0.1) | -0.01***<br>(-0.01, 0.00) | -0.01***<br>(-0.01, 0.00) | -0.01<br>(-0.03, 0.00) |
| Child recognition memory (controls only n=119)<sup>a,e</sup> | 0.00<br>(-0.01, 0.01) | 0.00<br>(-0.01, 0.02) | 0.01<br>(-0.01, 0.03) | -0.03<br>(-0.11, 0.04) |
| Child prenatal alcohol-related recognition memory deficits<sup>a,f,g</sup> | -0.02**<br>(-0.03, -0.01) | 0.02*<br>(0.00, 0.05) | 0.04*<br>(0.01, 0.07) | 0.17**<br>(0.06, 0.30) |
| Child FAS or PFAS diagnosis<sup>f,h</sup> | 1.03***<br>(1.01, 1.05) | 0.96**<br>(0.93, 0.98) | 0.96†<br>(0.92, 1.00) | 0.85*<br>(0.74, 0.99) |
| **<u>Child Ancestry (n = 280)</u>** |  |  |  |  |
| Child recognition memory (controls only (n=113); Z-scores)<sup>a,e</sup> | 0.00<br>(-0.01, 0.01) | 0.00<br>(-0.01, 0.02) | 0.01<br>(-0.02, 0.03) | 0.03<br>(-0.04, 0.10) |
| Child prenatal alcohol-related recognition memory deficits<sup>a,f,g</sup> | -0.02**<br>(-0.03, -0.01) | 0.03**<br>(0.01, 0.05) | 0.05**<br>(0.02, 0.08) | 0.17**<br>(0.05, 0.30) |
| Child FAS or PFAS diagnosis<sup>f,h</sup> | 1.03**<br>(1.01, 1.05) | 0.96**<br>(0.93, 0.99) | 0.95*<br>(0.91, 0.99) | 0.84*<br>(0.72, 0.99) |

†p ≤ 0.10; \*p ≤ 0.05; \*\*p ≤ 0.01; \*\*\*p ≤ 0.001.
AFR = African ancestry percentage; EAS = East ancestry percentage; EUR = European ancestry percentage; FAS = fetal alcohol syndrome; PFAS = partial FAS; SAS = South Asian ancestry percentage.
<sup>a</sup>Values are regression coefficients (95% confidence intervals) from linear models for each ancestry percentage for all maternal outcomes. For alcohol-related child recognition memory deficits, values are regression coefficients for ancestry percentage-by-prenatal alcohol exposure interaction terms from linear regression models.
<sup>b</sup>Model: Hollingshead socioeconomic score (46) = ancestry percentage + cohort.
<sup>c</sup>Model: maternal height (m) = ancestry percentage + cohort + socioeconomic status score. Missing for 2 mothers.
<sup>d</sup>Model: Average oz absolute alcohol/day across pregnancy (logged values) = ancestry percentage + cohort + socioeconomic status score.
<sup>e</sup>Z-scores were calculated within each cohort for recognition memory scores on the California Verbal Learning Test-Children's version (Pearson Clinical Assessments, USA) administered at age 10 years for Cohort 1 and on the Wide Range Assessment of Memory and Learning, 2<sup>nd</sup> Edition (Pearson Clinical Assessments, USA) administered at age 7 years for Cohort 2. Missing for 24 children.
<sup>f</sup>Excluding 32 children whose mothers (all heavy drinking women) received prenatal dietary choline supplementation as part of a pilot study.
<sup>g</sup>Model: Child recognition memory Z-score = ancestry percentage + average oz absolute alcohol/day across pregnancy (logged values) + (ancestry percentage)\*(average oz absolute alcohol/day across pregnancy (logged values)) + cohort + socioeconomic status score.
<sup>h</sup>Values are odds ratios (95% confidence intervals) for each ancestry percentage from logistic regression models. Model: Child diagnosis of FAS or PFAS = ancestry percentage + average oz absolute alcohol/day across pregnancy (logged values) + cohort + socioeconomic status score. Diagnoses made from at least one examination by expert fetal alcohol spectrum disorders dysmorphologists. Missing for 4 children.

Neither maternal nor child ancestry profiles were associated with child recognition memory among control children (all *p*’s :=0.376; **Table 4**).

Both maternal and child African ancestry percentages were associated with more severe alcohol-related recognition memory deficits in regression models examining ancestry-by-PAE interaction effects on child recognition memory (controlling for maternal age, socioeconomic status, and cohort; **Table 4**). Conversely, maternal and child European, South Asian, and East Asian ancestry percentages appeared to be protective.

In logistic regression models, after control for prenatal alcohol exposure, maternal age, socioeconomic status, and cohort, maternal African ancestry percentage was associated with higher odds of FAS/PFAS diagnoses (**Table 4**), whereas maternal European and East Asian ancestry percentages were related to lower odds of an FAS/PFAS diagnosis. For example, each 1% increase in maternal African ancestry percentage was associated with a 3% increase in odds of an FAS/PFAS diagnosis, and each 1% increase in East Asian ancestry percentage was associated with a 15% decrease in odds of these diagnoses. A trend was seen for lower odds with increasing maternal South Asian ancestry percentage. Child African ancestry percentage was also related to higher odds of FAS/PFAS diagnoses, whereas child European, Southern Asian, and East ancestry percentages were all related to lower odds of these diagnoses. For example, each 1% increase in child African ancestry percentage was associated with a 3% increase in odds of an FAS/PFAS diagnosis, and each 1% increase in East Asian ancestry percentage was associated with a 16% decrease in odds of these diagnoses.

## DISCUSSION

Utilizing genetic data from our two longitudinal birth cohorts of SACC mother-child dyads, we described the cohorts’ genetic admixture and documented a higher number of polymorphisms, especially rare, and high rates of homozygosity when compared to reference populations (i.e., European descent). We found that ancestry is associated with higher risks of multiple FASD-related outcomes, including worse cognitive performances. Nevertheless, among control dyads, no significant associations were observed between maternal or child ancestry and child cognition. Thus, we must emphasize that our findings show that ancestry is not associated with cognition but rather with more severe PAE-induced child cognitive deficits.

The genetic admixture of the SACC population represents a complex history of colonial slavery, migration, and labor practices, race-based state policies during the apartheid regime, and most recently, three decades of democracy. The SACC population is mainly comprised of descendants of farm workers in the Western Cape Province of South Africa, which was colonized by Portuguese and Dutch settlers in the 1700s (15, 40). Cape Town was a mandatory stopping point for all maritime trade between the Netherlands and all geographic regions east of Cape Town. Historic records show that, in the late 1700’s and early 1800’s, this included the trading of enslaved people by the Dutch East India Company, with a majority of enslaved people coming from East Africa (Bantu) and smaller numbers from Madagascar, the Bay of Bengal, the Southern coast of India, and the Indonesian Archipelago (40). Commercial trade also led to the migration into Cape Town of merchants and laborers from these regions, including the “free black’’ Chinese population, who formed ∼9% of the Cape Town population in the early 1800’s.

Much of the local Khoe-San population was also incorporated into the farm labor work force early in colonization of the region. The number of Bantu-speaking African peoples migrating into the Western Cape region from other parts of South Africa grew in the 1800’s and 1900’s. Under the apartheid regime, the government institutionalized the racial categories of White, Black, and Coloured; race category designations dictated who people could marry and where individuals could live, work, and freely pass. The Coloured designation included all people with mixed-ancestry, and government officials used subjective physical characteristics, such as skin color and hair, to determine an individual’s race designation, sometimes resulting in mismatches between self-identified and state-designated racial categories. During the transition to democracy in the early 1990s, new laws and a new constitution abolished prohibited discrimination based on race. Additionally, the Muslim-practicing population within the SACC is known as “Cape Malay” and has been thought to be somewhat ancestrally distinct, due to marriages being more likely to occur within religious groups and religious practices being different across the ancestral geographic regions of the SACC. Our findings confirmed this distinction, as Muslim-practicing dyads had notably higher South Asian ancestry proportions, lower African ancestry proportions, and slightly higher European and East Asian ancestry proportions.

Our findings are notably consistent with prior reports (15–19). de Wit et al. (16) estimated admixture in 959 SACC individuals, reporting ∼67% African, ∼23% European, and ∼10% Asian ancestry. In that study, reference populations from HapMap and HGDP were used but employed East Asian and Oceanic reference populations, which may explain differences between our findings and theirs. Daya et al. (15) employed genotype data from 733 SACC individuals and estimated ∼58% African, ∼18% European, ∼16% South Asian, and ∼9% East Asian ancestry. This study utilized a predominantly Indian (Gujarati) sample as a surrogate for South Asian ancestry population and Chinese and Japanese populations as surrogates for East Asian ancestry. Most recently, Choudhury et al. (19) conducted deep whole-genome sequencing on 8 SACC individuals and found higher proportions of South Asian and East Asian ancestry than in prior studies when using reference populations from Bangladesh and Malaysia, which may be more geographically appropriate given locations of the East Indian slave trade and Dutch trading outposts. Building on these studies, we used the same Bengali reference population as in Choudhury et al. (19) and reference populations from China and Southern regions of East Asia, which may give better estimates of ancestry from these regions. A key difference between our study and prior work is our overrepresentation of females in studying mother-child dyads, which is important given prior work demonstrating sex bias in the genetic admixture of the SACC population (41). Another notable difference between our cohorts and those of other studies is that most prior studies excluded Cape Malay individuals, who comprised ∼1/5 of our sample.

The genetic architecture of this population suggests that it may be optimal for identifying novel disease-associated genetic loci, given the evident enrichment for rare variants, which are more likely associated with clinical conditions as compared to common variants. For example, all of the monomorphic variants in the European reference population were non-monomorphic in our SACC cohorts; of these, 90% were common (MAF>1%). Mean homozygosity in this population was also quite high (6%); despite the fact that African-descended populations exhibit lower rates of homozygosity (as confirmed in our analyses), homozygous genetic segments of African ancestral origin have been shown to have greater deleterious variation (42). This finding is indeed replicated in our SACC cohort. The general pattern of high minor allele frequencies, with high proportions of rare variants and high rates of homozygosity is likely to boost power to detect associations between genetic loci and phenotypic outcomes and/or diseases. Furthermore, relative to more homogeneous populations and cohorts with diversity *between* individuals, high genetic admixture *within* individuals, as is the case of our cohorts, increases the likelihood that prioritized genetic loci are generalizable to other worldwide populations.

In our sample, in which roughly half of the children had heavy PAE, we found that ancestry pattern was associated with altered odds of a diagnosis of FAS or PFAS, as well as altered severity of PAE-related recognition memory deficits, indicating that ancestry-specific signals may help explain underlying FASD pathophysiology and/or phenotypic variability and may ultimately help to identify to risk-modifying loci in the mother or child. In our cohorts, a general pattern was observed, where maternal and child African ancestry was associated with increased risk of FASD outcomes, and conversely European, South Asian, and East Asian ancestries were found to be protective. It is important to note that since ancestry proportions are interdependent and add up to 100%, increased African ancestry correlates with decreased ancestry from other subgroups. Thus, our findings do not differentiate between whether higher African proportion is truly risk-conferring, independent of higher European, South Asian, and/or East Asian ancestry proportions or, conversely, whether higher European, South Asian, and/or East Asian ancestry proportions are protective, independent of African ancestry proportion. Future studies with admixture mapping are needed to identify whether ancestry-specific genetic segments are associated with altered risk of FASD. Admixture mapping identifies associations between disease outcomes and ancestry-specific genetic segments, which are much more infrequent than SNPs or genes; disease-associated genetic segments can then be further explored to determine which loci drive the disease associations (12). Although our findings were notably independent of socioeconomic status scores, residual confounding by socioenvironmental factors is likely still present in our analyses given worldwide discrimination based on characteristics associated with race and ethnicity. In South Africa, state laws discriminating against people based on skin color and hair were present during some of our cohorts participants’ lifetimes, and ancestry profiles were associated with different socioeconomic status scores despite participants coming from the same communities. Additionally, being Cape Malay, which may be associated with differential socioenvironmental factors, including diet, early education, and drinking patterns, was not related to any maternal or child outcome.

The SACC cohorts’ genetic features create unique avenues for understanding the molecular and cellular basis of FASD through experimental models. For example, this study enables further hypotheses for investigating the cellular, molecular, and behavioral basis of FASD through the integration of developmental and molecular biology perspectives. Research on how specific genetic factors influence fetal development could uncover critical periods of vulnerability to alcohol exposure, enhancing our understanding of FASD susceptibility. Additionally, the study of gene-environment interactions could reveal how genetic predispositions and alcohol consumption interplay to affect fetal development, offering deeper insights into FASD pathogenesis and potential intervention points. The functional characterization of genetic variants associated with FASD could elucidate the molecular mechanisms. Such characterization could involve assessing how these variants impact gene expression, protein function, and cellular pathways. Experimental assay systems such as animal models carrying genetic variants linked to FASD to observe developmental and molecular consequences in vivo could significantly contribute to this perspective. Cellular models derived from induced pluripotent stem cell-derived cell types of individuals with FASD could help characterize the effects of specific genetic patterns on cell differentiation and development under alcohol exposure. Finally, pharmacogenomics studies could pave the way for personalized medicine by understanding how genetic variations influence responses to therapeutic interventions for FASD.

This study had limitations common to other genetic epidemiology studies. Our study recruitment strategy was designed to overrepresent heavy-drinking pregnant women; thus the cohorts may not be representative of the general SACC population in South Africa. Furthermore, the cohorts were recruited from a predominantly peri-urban area and may not be fully representative of SACC individuals across South Africa. Although the gene array (MegaEX) used is among the most accurate for multi-ethnic populations, genotyping errors are higher in genetically admixed populations than in others (43).

Furthermore, recent analysis of imputation error rates in diverse global populations revealed that imputation errors are exceedingly high in the African San and East Asian populations (especially in the Indonesian Archipelago) (44), which comprise ∼40% and 5% of the genetic admixture in our sample, respectively. These issues highlight the need for deep whole genomic sequencing in the SACC population for future studies. Of note, our associations of maternal ancestry profiles with maternal height and % homozygosity are consistent with what is expected, which provides evidence of validity of our ancestry estimates despite potential genotyping/imputation errors. Although PAE estimates were assessed by self-report, and thus subject to recall error, interviews were conducted prenatally, and we have previously demonstrated the predictive validity of the timeline follow-back interview used (26). Errors in phenotype assessments are also a concern in genetic epidemiology studies, but all participants were examined by a group of world-renowned FASD dysmorphologists and each diagnosis was confirmed by at least two dysmorphologists (45). Furthermore, the pattern of interaction effects between ancestry profiles and PAE were generally consistent across multiple independently measured neurobehavioral outcomes.

In summary, we documented genetic admixture consistent with prior reports although with higher proportion of South Asian and East Asian ancestries. We also confirmed marked enrichment for rare variants, and a high degree of homozygosity. These features indicate that the SACC population may be ideal for genetic disease-association studies when compared to other populations. The associations between ancestry and FASD-related outcomes support this premise. Future studies are needed in larger cohorts; considering the array and imputation error rates among the SACC’s ancestral San and East Asian populations, whole genome sequencing would greatly improve the quality of findings in such studies.

## ACKNOWLEDGMENTS

We thank our UCT and Wayne State University research staff including Maggie September, Beverley Arendse, Patricia O’Leary, and Patricia Solomon for their work on subject recruitment and maintenance; and Renee Sun, for her assistance with data processing. We thank Catherine O’Leary, Nadine Lindinger, and Natalia Berghoff for their work on child neurobehavioral assessments. We also thank the dysmorphologists H. Eugene Hoyme, Luther Robinson, and Nathaniel Khaole for their work in examining the children. We thank Yula Ma (Mt. Sinai) and Qian Li (Columbia University) for their work on DNA isolation. We also thank Susan Fawcus, MD, Head of Department of Obstetrics, Mowbray Maternity Hospital; the nursing and records department staff at the Hanover Park and Retreat Midwife Obstetric Units, Mowbray Maternity Hospital, Somerset Hospital, and Groote Schuur Hospital, where the mothers were recruited and the infants were born. Lastly, we extend our deep appreciation to the Cape Town study participants for their contributions to this study.

## CONFLICTS OF INTEREST

The authors declare no conflict of interest.

## FUNDING

This research was funded by NIH/NIAAA (R01AA016781, R01AA027916, R01AA023503, K23AA020516, U01AA014790), NIH/NIA (U01AG081817, RF1AG082009, U19AG074865), and the Lycaki-Young Fund (State of Michigan). The funding sources had no role in study design; in the collection, analysis and interpretation of data; in the writing of the report; and in the decision to submit the article for publication.

## Data Availability Statement

De-identified, individual participant data that underlie the results reported in this article and the study protocol, statistical analysis plan, and analytic code will be available for sharing to journal editors for any reason either before or after publication for checking and to researchers who provide a methodologically sound proposal, as determined by the authors of this article. Proposals from interested parties should be directed to Sandra W. Jacobson, PhD.

Data will be stored in a data repository at Wayne State University and transmitted electronically in encrypted form to requestors. Data requestors will need to sign a data access agreement prior to access.

## Author Contributions

Conceptualization, T.A.H., J.L.J, C.K., R.C.C., S.W.J., G.T., S.H.Z.; methodology, J.L.J, R.C.C., S.W.J., G.T.; formal analysis, R.C.C., G.T., Z.Y.; resources, E.M.M., J.L.J, R.C.C., S.W.J., S.H.Z.; data curation, N.C.D., H.E.H., J.L.J, R.C.C., S.W.J., G.T., S.H.Z., Z.Y.; writing—original draft preparation, R.C.C., G.T., Z.Y.; writing—review and editing, T.A.H., H.E.H., J.L.J, S.W.J., C.K., S.H.Z.; funding acquisition, E.M.M., J.L.J, R.C.C., S.W.J., G.T., S.H.Z.; All authors have read and agreed to the published version of the manuscript.

## Institutional Review Board Statement

The study was conducted in accordance with the Declaration of Helsinki, and approved by the Institutional Review Boards/Ethics Committees of Wayne State University (026708B3F, approved August 2011 and annually thereafter), University of Cape Town Faculty of Health Sciences, and Columbia University Irving Medical Center.

## Informed Consent Statement

Informed consent was obtained from all mothers involved in the study.

## Notes

### Competing Interest Statement

The authors have declared no competing interest.

### Summary of Updates

A formatting error to Table 1 that occurred during PDF file conversion has been fixed. No content has been changed.

