## Supplemental Materials for "Genetic admixture predictors of fetal alcohol spectrum disorders (FASD) in the South African Cape Coloured population"

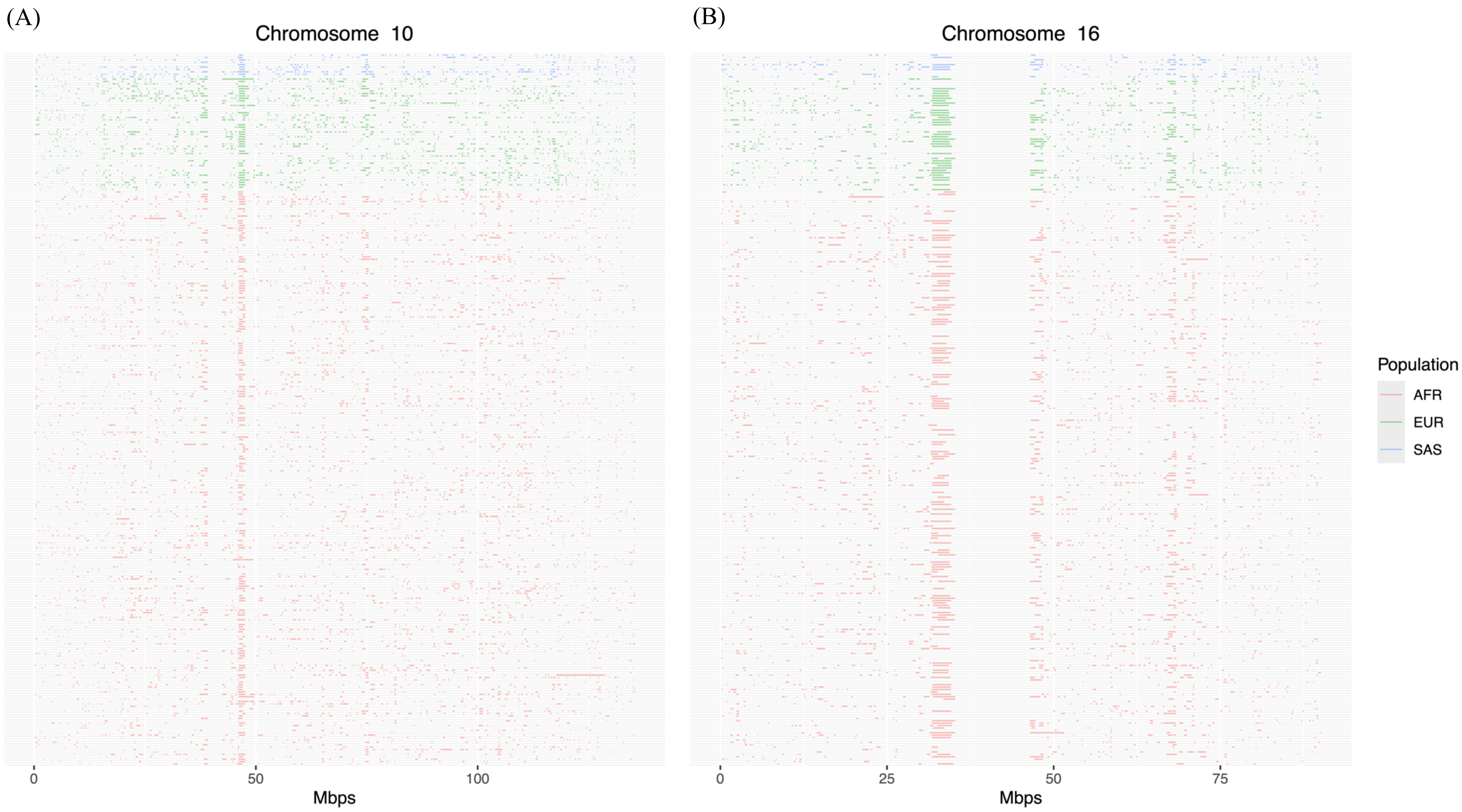


**Figure S1.** Runs of homozygosity (ROHs) with high overlap, color-coded by ancestral origin, among 296 unrelated mothers in chromosomes 10 (A) and 16 (B). Ancestry designation was made if >80% of variants within the ROH traced back to a given ancestry.

**Table S1**. Average ancestral percentage (%) of samples by populations[a]

| | | K | 1 | 2 | 3 | 4 | 5 | 6 |
|---|---|---|---|---|---|---|---|---|
| | | Study sample (SACC) | 14.5 | 42.7 | 15.7 | 18.2 | 1.9 | 6.9 |
| REFERENCE POPULATIONS | AFRICAN | Yoruba | 99.3 | 0.7 | 0 | 0 | 0 | 0 |
| | | Mandenka | 99.6 | 0 | 0 | 0.3 | 0 | 0 |
| | | Gambian | 99.7 | 0 | 0 | 0.3 | 0 | 0 |
| | | Bantu South Africa | 63 | 37 | 0 | 0 | 0 | 0 |
| | | Bantu Kenya | 72.6 | 26 | 0.2 | 1.2 | 0 | 0 |
| | | San | 0 | 100 | 0 | 0 | 0 | 0 |
| | EUROPEAN | French | 0.1 | 0 | 0.1 | 99.8 | 0 | 0 |
| | | British | 0 | 0 | 0 | 100 | 0 | 0 |
| | | Italian | 0 | 0 | 1.9 | 98.1 | 0 | 0 |
| | | Northern European | 0 | 0 | 0 | 100 | 0 | 0 |
| | SOUTH ASIANS | Bengali | 0 | 0 | 94.3 | 1.9 | 2.4 | 1.4 |
| | EAST ASIANS | Chinese South | 0 | 0 | 0 | 0 | 79 | 21 |
| | | Chinese North | 0 | 0 | 0.5 | 0.2 | 87.9 | 11.5 |
| | | Cambodian | 0 | 0 | 10.2 | 0 | 23.8 | 66 |
| | | Vietnamese (Kinh) | 0 | 0 | 0.1 | 0.1 | 14 | 85.7 |
| | | Lahu | 0 | 0 | 0 | 0 | 34.3 | 65.7 |

[a]Results were computed by ADMIXTURE (1) with the number of ancestries set at 6 (K=6) using uncorrelated genetic markers in 352 unrelated SACC individuals and HGDP and 1000G African, European, South Asian, and East Asian reference populations.

**Table S2**. Ancestry proportion comparisons between Cape Malay and all other Cape Coloured participants[a]

| | Cape Malay | | All other Cape Coloured | | |
|---|---|---|---|---|---|
| | *n* | *M* (SD) | *n* | *M* (SD) | $p^b$ |
| Maternal Ancestry | | | | | |
| % African | 63 | 42.9 (17.2) | 218 | 54.9 (19.4) | <0.001 |
| % European | 63 | 28.0 (11.1) | 218 | 24.9 (13.0) | 0.053 |
| % South Asian | 63 | 23.5 (8.9) | 218 | 16.3 (8.5) | <0.001 |
| % East Asian | 63 | 5.8 (2.9) | 218 | 4.0 (2.5) | <0.001 |
| Child Ancestry | | | | | |
| % African | 55 | 41.2 (12.7) | 202 | 53.2 (18.0) | <0.001 |
| % European | 56 | 28.9 (9.0) | 202 | 25.3 (11.2) | 0.013 |
| % South Asian | 56 | 24.4 (6.8) | 202 | 17.3 (8.0) | <0.001 |
| % East Asian | 56 | 5.5 (2.5) | 202 | 4.2 (2.4) | <0.001 |

[a]Cape Malay status missing for 28 mother-child dyads.
[b]From independent samples *t*-tests

**Table S3**. Correlations between 4-group ancestry[a] and runs of homozygosity among 296 unrelated mothers[b]

| **Ancestry** | **Homozygosity** |
|---|---|
| % African | -0.89*** |
| % European | 0.78*** |
| % South Asian | 0.70*** |
| % East Asian | 0.49*** |

***$p < .001$.
[a]Estimated with ELAI (2) method.
[b]Values are Pearson *r*.

**Table S4**. Prevalence of deleterious variants in the 10 most overlapping runs of homozygosity among 296 unrelated mothers

| Local Ancestry | # of mothers with ROH | Total # of deleterious variants | Poisson *p* |
|---|---|---|---|
| AFR | 27 | 19 | 0.023 |
| non-AFR | 64 | 21 | |
